# Precision Diagnostic Approach to Predict 5-year Risk for Microvascular Complications in Type 1 Diabetes

**DOI:** 10.1101/2021.09.28.21264161

**Authors:** Naba Al-Sari, Svetlana Kutuzova, Tommi Suvitaival, Peter Henriksen, Flemming Pociot, Peter Rossing, Douglas McCloskey, Cristina Legido-Quigley

**Affiliations:** Steno Diabetes Center Copenhagen, Denmark; Novo Nordisk Foundation Center for Biosustainability, Technical University of Denmark; Dept. of Clinical Medicine, University of Copenhagen, Denmark; Institute of Pharmaceutical Science, King’s College London, UK

**Keywords:** Diabetes Complications, Diabetic Kidney Disease, Diabetic Retinopathy, Machine Learning, Microvascular Complications

## Abstract

**Background:** Individuals with long standing diabetes duration can experience damage to small microvascular blood vessels leading to diabetes complications (DCs) and increased mortality. Precision diagnostic tailors a diagnosis to an individual by using biomedical information. Blood small molecule profiling coupled with machine learning (ML) can facilitate the goals of precision diagnostics, including earlier diagnosis and individualized risk scoring.

**Methods:** Using data in a cohort of 537 adults with type 1 diabetes (T1D) we predicted five-year progression to DCs. Prediction models were computed first with clinical risk factors at baseline and then with clinical risk factors and blood-derived molecular data at baseline. Progression of diabetic kidney disease and diabetic retinopathy were predicted in two complication-specific models.

**Findings:** The model predicts the progression to diabetic kidney disease with accuracy: 0.96±0.25 and 0.96±0.06 area under curve, AUC, with clinical measurements and with small molecule predictors respectively and highlighted main predictors to be albuminuria, glomerular filtration rate, retinopathy status at baseline, sugar derivatives and ketones. For diabetic retinopathy, AUC 0.75±0.14 and 0.79±0.16 with clinical measurements and with small molecule predictors respectively and highlighted key predictors, albuminuria, glomerular filtration rate and retinopathy status at baseline. Individual risk scores were built to visualize results.

**Interpretation:** With further validation ML tools could facilitate the implementation of precision diagnosis in the clinic. It is envisaged that patients could be screened for complications, before these occur, thus preserving healthy life-years for persons with diabetes.

**Funding:** This study has been financially supported by Novo Nordisk Foundation grant NNF14OC0013659.

*Research in context:* 

*Evidence before this study:* Microvascular diabetes complications (DCs), such as diabetic kidney disease (DKD) and diabetic retinopathy (DR), lead to increased mortality, blindness, kidney failure and overall decreased quality of life in individuals with diabetes.

*Added value of this study:* We have developed four algorithms based on traditional risk factors alone and risk factors with metabolomic and lipidomic data. Two accurately predicted the future progression in individuals with type 1 diabetes. The top predictors chosen by the algorithm were hemoglobin A_1c_, albuminuria and retinopathy status at baseline for diabetic retinopathy. The most promising models predicted future diabetic kidney disease in which albuminuria, eGFR, retinopathy status at baseline were complemented with a number of ketones and sugar derivatives.

*Implications of all the available evidence:* With further validation in several cohorts, the prediction models presented here have the potential for early diagnosis of CKD in persons with diabetes, thus enabling appropriate decisions on the available therapies. It is envisaged that these algorithms will be trialed and support clinicians with precision diagnosis and treatments.

## 1. Introduction

Devastating microvascular diabetes complications (DCs), such as diabetic nephropathy (DN) and diabetic retinopathy (DR), lead to increased mortality, blindness, kidney failure and overall decreased quality of life in individuals with diabetes (1,2). Systemic high glucose levels result in damage in the cells of the capillary endothelium of the retina and in the cells of the mesangial in the glomerulus. Thus, hyperglycemia is the most important known predictor of the pathogenesis of these two complications in type 1 diabetes (3). Glomerular filtration rate (GFR) and the urinary albumin excretion rate, which themselves are measures of DN, are also major predictors of further progression of DN (4). Although clinical risk factors and glycemic control can be good predictors of the development of microvascular complications, they are not necessarily informative at the early stages of disease. Hence, there is a need for technology that can exploit hidden risk patterns and molecular dynamics, thus achieving accurate prediction of DCs.

Metabolomics and lipidomics are snap-shots of metabolism and can be applied to the study of diabetes to obtain a comprehensive molecular profile (5). Over the past decade, omics technologies have shown the potential to personalize patient care in a way that was previously unthinkable (6-8). Thus, by combining already well-known clinical risks factors together with a broad omics panel, we aim to study the biological dynamics during progression to DR and DN.

Machine learning (ML) algorithms learn descriptive patterns from large amounts of data. Hence, the application of this technology can support clinical decisions and is one of the areas where artificial intelligence has had the most impact in the recent years (9-10). ML can empower healthcare professionals, and to date it has been applied effectively to predict the risk of heart failure and retinopathy in diabetes (11,12). Significant strides are being made towards using ML algorithms to predict other conditions. In the case of diabetic kidney disease (DKD), extensive research has been carried out to find predictive biomarkers for future end-stage kidney disease. However, to the knowledge of the authors, no single study exists, which employs ML to predict progression of eGFR decline in type 1 diabetes (13). In the case of DR complications, on the other hand, hundreds of publications and patents with highly predictive approaches have recently been reported and filed, including deep learning-based image analysis of retinal images (12).

In a large and well-characterized type 1 diabetes cohort from Steno Diabetes Center Copenhagen (SDCC), we sought to develop easily interpretable and accurate prognostic risk prediction models for DCs. To this end, we apply ML with clinical data combined with two sets of omics data to predict DC progression in follow-up data. In this study, we hypothesize that (1) ML can be used for prediction of future complications in type 1 diabetes using standard clinical risk factors; and (2) combining blood-based metabolic phenotyping and clinical data will improve the prediction by modeling the dynamics between risk factors and molecular metabolism. The ultimate aim of this study is to design a personalized risk prediction tool for DCs that can be applied in clinical practice.

## 2. Methods

### 2.1 Study design and Participants

This study is based on a cohort of 648 adults with type 1 diabetes followed at SDCC and previously described by Theilade, et al. (14). As the present study focuses on prediction of progression, any participants with missing follow-up data on DCs were excluded from the analysis. Thus, metabolomics and lipidomics data along with follow-up information on DKD and retinopathy status were available for 537 participants. Advanced DCs at baseline such as macroalbuminuria, and severe retinopathy (proliferative or blind) were excluded, leaving 385 participants with mild or no DCs at the baseline assessment.

The study was performed in compliance with the Declaration of Helsinki and was approved by the ethics committee for the Capital Region of Denmark (Hillerød, Denmark). All participants have given a written consent.

### 2.2 Baseline Clinical Measurements

A detailed description of clinical measurements has previously been reported (15-17). HbA_1c_, serum creatinine, plasma cholesterol, and triglycerides were measured using standardized methods from venous samples. Albuminuria was subdivided by stages (normo-, micro-, and macroalbuminuria, using 30 and 300 mg/g creatinine or mg/24 hours as cut offs). Decline in eGFR was defined as the first occurrence of ≥30% decrease from baseline, as proposed by Coresh et al. (18). Retinopathy status was assessed at SDCC as no retinopathy, mild non-proliferative retinopathy, moderate non-proliferative retinopathy, proliferative retinopathy, proliferative retinopathy with fibrosis, and blind. Previous cardiovascular disease (CVD) was defined as any previous event of ischemic heart disease, ischemic stroke, heart failure, and peripheral artery disease. Information on medication was collected from electronic medical records. Following categories were applied: use of lipid-lowering treatment (yes/no), antihypertensive treatment (yes/no) and current smoking (yes/no).

### 2.3 Metabolic phenotyping and preprocessing

Metabolite and lipid concentrations were measured in plasma samples using untargeted ultra-high-performance liquid chromatography coupled to mass spectrometry (UHPLC-MS) and two-dimensional gas chromatography coupled to time-of-flight mass spectrometry (GC×GC-TOFMS) as previously described (16,17,19-21). Global metabolomics based on GC×GC-TOF-MS, covers small molecules such as sugars, free fatty acids and amino acids. Global lipidomics based on UHPLC-MS covers molecular lipid species, such as neutral lipids, sphingolipids and phospholipids. Raw GC×GC-TOF-MS and UHPLC-MS data were processed with ChromaTOF (LECO; Saint Joseph; USA) and MZmine 2, respectively. Finally, data from each platforms were post-processed in R by batch-correction, truncation of outliers, and imputation of missing values, as described previously. The final data sets of metabolite and lipid species consisted of the measured levels of identified and unidentified compounds. Inclusion of the complete data from the platforms was used to acquire an unbiased global metabolic phenotype. Finally, features with a very high mutual correlation (Pearson correlation coefficient larger than 0.85) were removed, thus leaving one feature from each tight feature group as a nonredundant predictor.

### 2.4 Machine learning method

#### 2.4.1 Data and model design

Random Forest (RF) models were applied to predict future risk of progression of DCs (22). We evaluated three scenarios with participants divided into two groups: first, non-progressors (persons with mild complication not advancing to another stage of the complication; n=195), and progressors (n=190), including both the DR and DKD progressors; second, only progression to DR (mild, moderate or severe) predicted for 193 non-progressors and 111 progressors; and third, only progression to DKD (≥30% decline in eGFR) predicted for 193 non-progressors and 79 progressors.

The RF classifier was employed to predict whether the participant will during the follow-up progress to at least one of the complications. For each of the models, two sets of features were evaluated: 1) clinical variables only (17 measures), and 2) blood small molecule data (965 molecular features) along with the clinical variables.

Clinical variables with no predictive power were excluded for improved performance. Unidentified compounds that were picked by the ML algorithm (as described next) were further investigated to acquire putative identities by manually comparing the retention time (RT), mass-to-charge ratio (m/z) and fragmentation pattern with spectral libraries. All prediction models were developed using SciKit-learn (23) in Python (v3.7.1). RF scalable visualizations (decision trees) for interpretation are created with ‘pybaobabdt’ library (24).

#### 2.4.2 Model validation

The models were trained by splitting the data into training and testing datasets through k-fold cross validation (k=5). The number of Decision Trees in the ensemble was set to 500, all features were included in each split without *a priori* feature selection and a panel of features was then selected by the model. Non-progressors and progressors were divided randomly into a training set (80%) used to build the RF models and an unseen validation data set (20%) used to validate the model performance. The main predictors and features of importance were selected using the maximal mean absolute SHapley Additive exPlanation (SHAP) algorithm. This method selects features while ensuring the top performance model is obtained (25). For each panel of predictors, the performance was calculated for each round on mean AUROC values from which the optimal number of features was selected. The models were further tested for performance stability using Monte Carlo (MC) simulation consisting of 50 iterations (26,27). Model performance was evaluated with the following metrics: AUROC, prediction accuracy, precision, recall (sensitivity), and F-score. Prediction performance was assessed at the class decision threshold of 0.50. To illustrate the applicability of the algorithm for personalized medicine, the results from two individuals using clinical risk factors were portrayed (Figure 2.G).

## 3. Results

A graphical representation of the study design and ML specifications is shown in Figure 1. Participants’ baseline characteristics are given in Table 1.

**Table 1.**
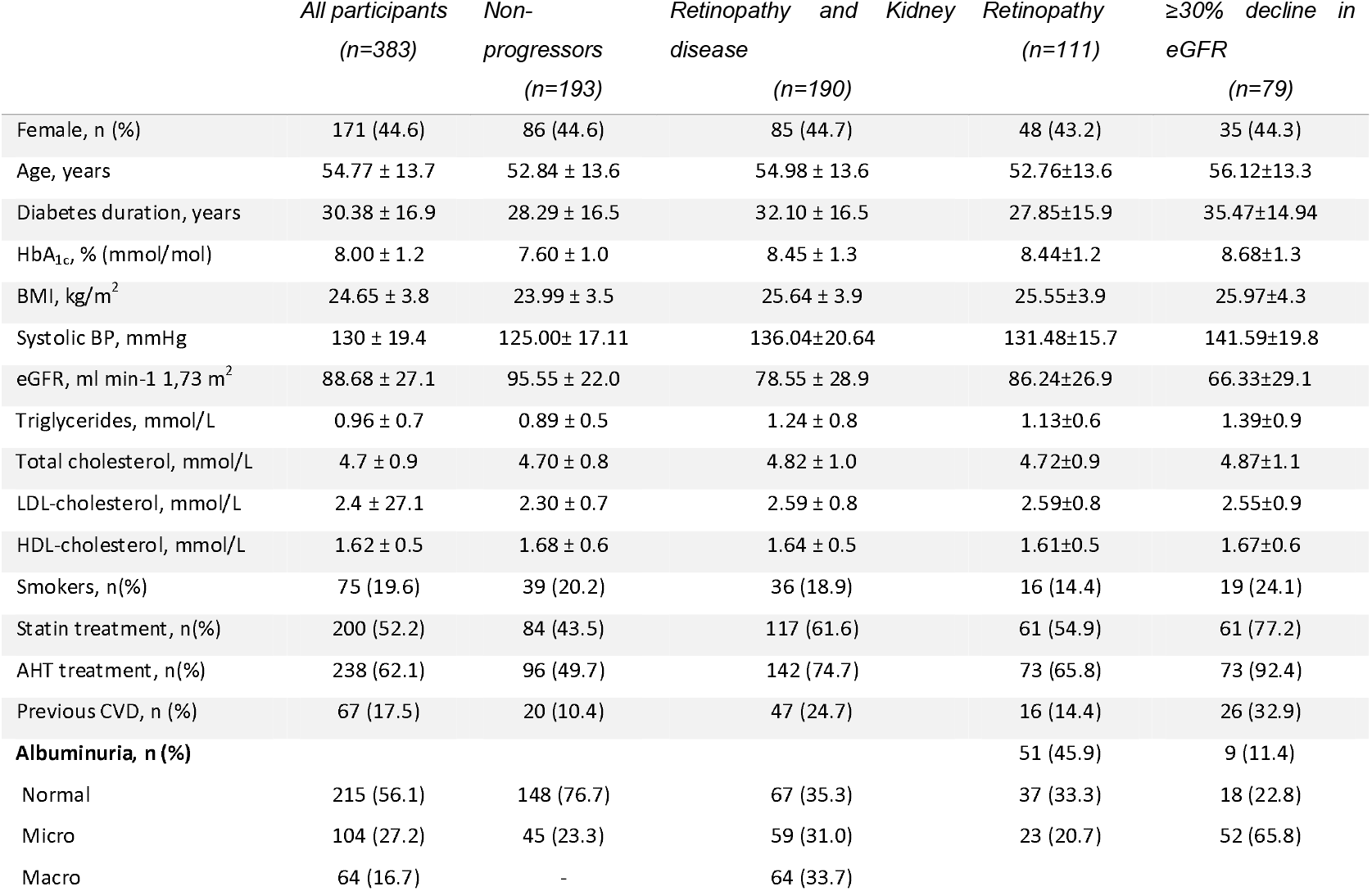
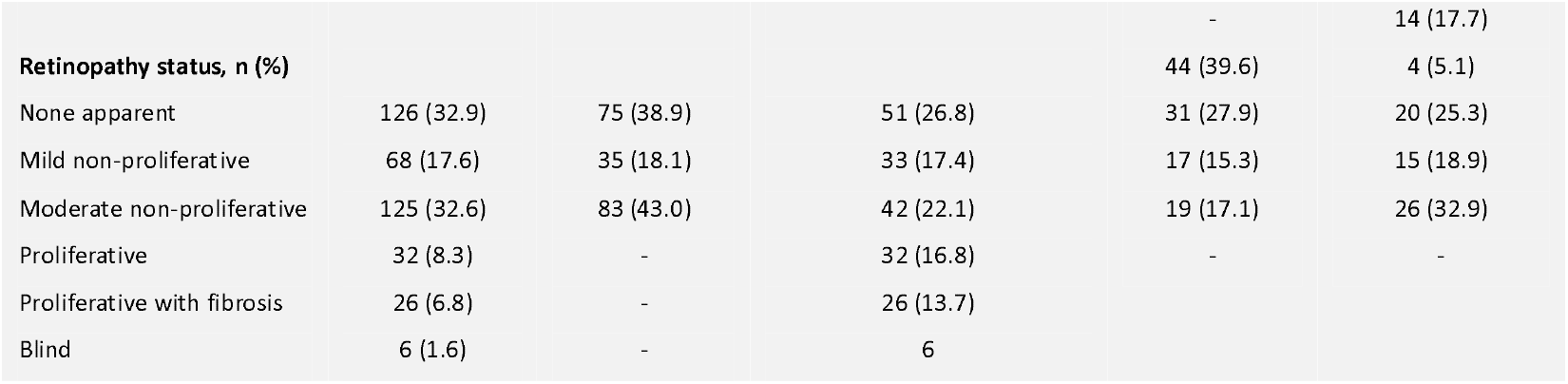
Comparison of baseline clinical characteristics of combined diabetes complications, diabetic retinopathy, and diabetic kidney disease. Data are n (%, rounded) and mean ±SD.

|  | *All participants*<br>*(n=383)* | *Non-*<br>*progressors*<br>*(n=193)* | *Retinopathy and Kidney*<br>*disease*<br>*(n=190)* | *Retinopathy*<br>*(n=111)* | *$\geq 30\%$ decline in*<br>*eGFR*<br>*(n=79)* |
| --- | --- | --- | --- | --- | --- |
| Female, n (%) | 171 (44.6) | 86 (44.6) | 85 (44.7) | 48 (43.2) | 35 (44.3) |
| Age, years | 54.77 ± 13.7 | 52.84 ± 13.6 | 54.98 ± 13.6 | 52.76±13.6 | 56.12±13.3 |
| Diabetes duration, years | 30.38 ± 16.9 | 28.29 ± 16.5 | 32.10 ± 16.5 | 27.85±15.9 | 35.47±14.94 |
| HbA<sub>1c</sub>, % (mmol/mol) | 8.00 ± 1.2 | 7.60 ± 1.0 | 8.45 ± 1.3 | 8.44±1.2 | 8.68±1.3 |
| BMI, kg/m<sup>2</sup> | 24.65 ± 3.8 | 23.99 ± 3.5 | 25.64 ± 3.9 | 25.55±3.9 | 25.97±4.3 |
| Systolic BP, mmHg | 130 ± 19.4 | 125.00± 17.11 | 136.04±20.64 | 131.48±15.7 | 141.59±19.8 |
| eGFR, ml min<sup>-1</sup> 1.73 m<sup>2</sup> | 88.68 ± 27.1 | 95.55 ± 22.0 | 78.55 ± 28.9 | 86.24±26.9 | 66.33±29.1 |
| Triglycerides, mmol/L | 0.96 ± 0.7 | 0.89 ± 0.5 | 1.24 ± 0.8 | 1.13±0.6 | 1.39±0.9 |
| Total cholesterol, mmol/L | 4.7 ± 0.9 | 4.70 ± 0.8 | 4.82 ± 1.0 | 4.72±0.9 | 4.87±1.1 |
| LDL-cholesterol, mmol/L | 2.4 ± 27.1 | 2.30 ± 0.7 | 2.59 ± 0.8 | 2.59±0.8 | 2.55±0.9 |
| HDL-cholesterol, mmol/L | 1.62 ± 0.5 | 1.68 ± 0.6 | 1.64 ± 0.5 | 1.61±0.5 | 1.67±0.6 |
| Smokers, n(%) | 75 (19.6) | 39 (20.2) | 36 (18.9) | 16 (14.4) | 19 (24.1) |
| Statin treatment, n(%) | 200 (52.2) | 84 (43.5) | 117 (61.6) | 61 (54.9) | 61 (77.2) |
| AHT treatment, n(%) | 238 (62.1) | 96 (49.7) | 142 (74.7) | 73 (65.8) | 73 (92.4) |
| Previous CVD, n (%) | 67 (17.5) | 20 (10.4) | 47 (24.7) | 16 (14.4) | 26 (32.9) |
| **Albuminuria, n (%)** |  |  |  | 51 (45.9) | 9 (11.4) |
| Normal | 215 (56.1) | 148 (76.7) | 67 (35.3) | 37 (33.3) | 18 (22.8) |
| Micro | 104 (27.2) | 45 (23.3) | 59 (31.0) | 23 (20.7) | 52 (65.8) |
| Macro | 64 (16.7) | - | 64 (33.7) |  |  |
|  |  |  |  | - | 14 (17.7) |
| **Retinopathy status, n (%)** |  |  |  | 44 (39.6) | 4 (5.1) |
| None apparent | 126 (32.9) | 75 (38.9) | 51 (26.8) | 31 (27.9) | 20 (25.3) |
| Mild non-proliferative | 68 (17.6) | 35 (18.1) | 33 (17.4) | 17 (15.3) | 15 (18.9) |
| Moderate non-proliferative | 125 (32.6) | 83 (43.0) | 42 (22.1) | 19 (17.1) | 26 (32.9) |
| Proliferative | 32 (8.3) | - | 32 (16.8) | - | - |
| Proliferative with fibrosis | 26 (6.8) | - | 26 (13.7) |  |  |
| Blind | 6 (1.6) | - | 6 |  |  |

**Fig. 1.**
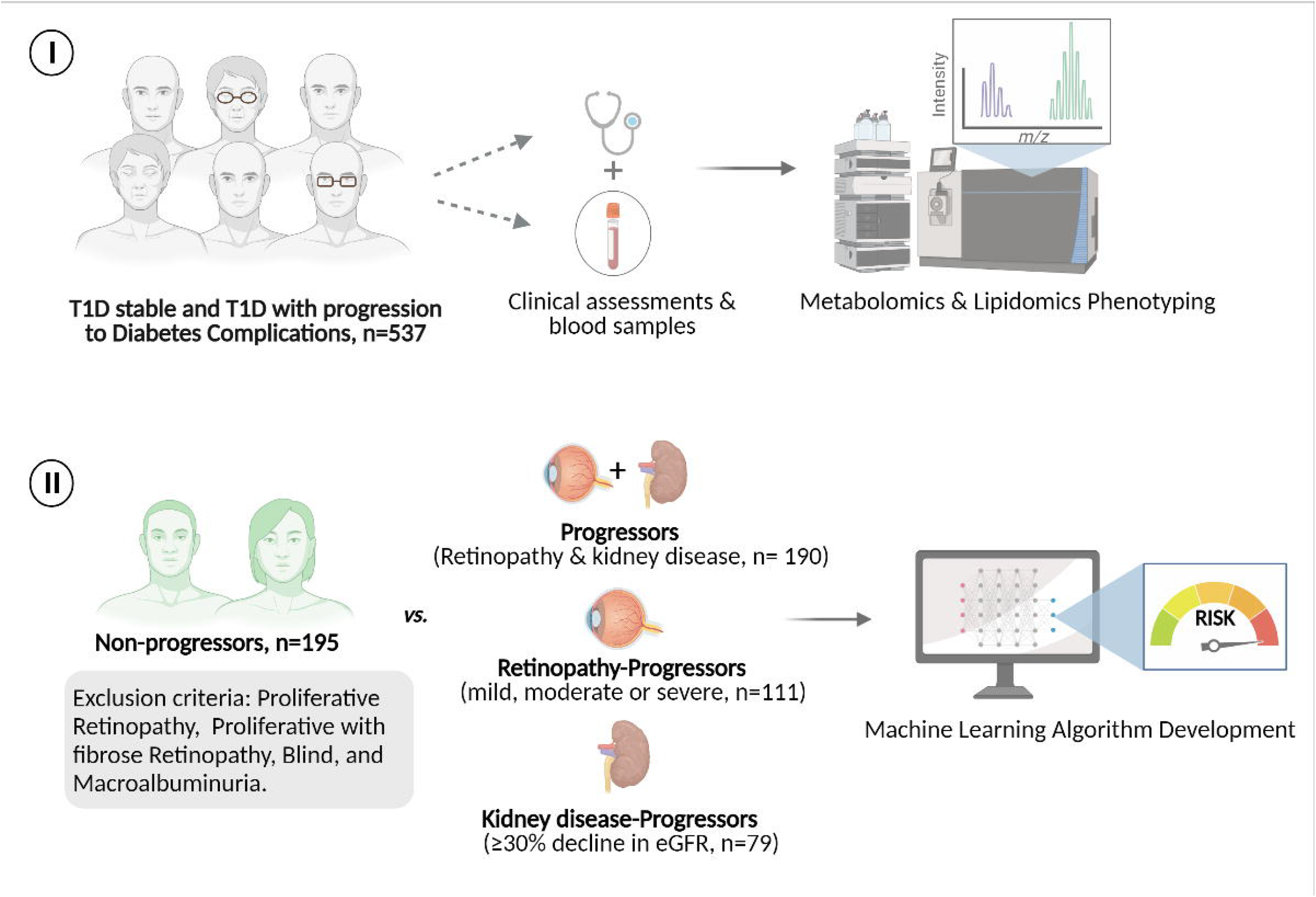
Graphical representation of study design and machine learning implication. Baseline clinical data and plasma samples (for metabolomics and lipidomics analysis) were collected from 537 individuals with type 1 diabetes (**I**). Participants were classified into two groups: type 1 diabetes stable (n=195) or type 1 diabetes with progression to diabetes complications (n=190). Progression of combined diabetes complications, diabetic kidney disease (≥30% decline in eGFR; n=79), and diabetic retinopathy (mild, moderate or severe; n=111) were predicted (**II**). Median follow-up was 5.4 years.

### 3.1 Baseline characteristics of the cohort

The baseline characteristics of the included individuals were as follows: mean ± SD: age of 54.8 ± 13.7 years, a median diabetes duration of 30.4 ± 16.9 years and 171 (45 %) women (**Table 1**). Overall, 215 (56 %) had normo-albuminuria at baseline, 104 (27 %) and 64 (17 %) had microalbuminuria and macroalbuminuria, respectively. At baseline eGFR was 88.8 ± 27.1 ml min^-1^ 1.73 m^-2^. During follow-up, 79 participants experienced a ≥ 30% decline in eGFR, and 111 individuals progressed in the DR stage. The majority (62 %) were on antihypertensive treatment (AHT) and statin (52 %) treatment. Median follow-up time was 5.4 years.

### 3.2 Metabolic Phenotyping

Using the two untargeted analytical platforms for metabolites, a total of 702 lipid species and 263 metabolites were measured, respectively, from 385 plasma samples. All 965 non-redundant omics features were included in the development of the ML algorithms (see methods, ‘Modeling design’).

Out of the omics features included, 14 omics features appeared as predictors of importance in the models with clinical and omics (described in the next subsection). Six of the selected omics features were known metabolites: ketone bodies (2,4-and 3,4-dihydroxybutanoic acids) and four sugar derivates (ribitol, ribonic acid, myo-inositol, and meso-erythrinitol).

Further two features were known lipid species: a saturated ceramide Cer(d42:0) and a monounsaturated sphingomyelin SM(d30:1). The last six features were putatively identified. Based on RT and Golm Metabolome Database (28), ‘M_68’ is a small metabolite with more than one hydroxyl group, thus likely a sugar. ‘M_76’ is indicatively a large carboxylic acid. Based on m/z values from the unknown lipid species and according to the LIPID MAPS database (29), ‘L_195’ and ‘L_168’ are putative ceramides. L_103’ is a phosphatidylserine or a phosphatidylinositol. ‘L_439’ could not be identified.

### 3.3 Risk Prediction Models

Overall, the predictive performance of all models using traditional risk factors showed excellent and robust predictive performance for future progression to DCs (**Figure 2**). Moreover, combining metabolic phenotyping and clinical variables improved the prediction performance (**Figure 3**). The importances and contributions of the top-features in the models are shown in the SHAP summary plots. Next, we will describe the models in detail.

**Fig. 2.**
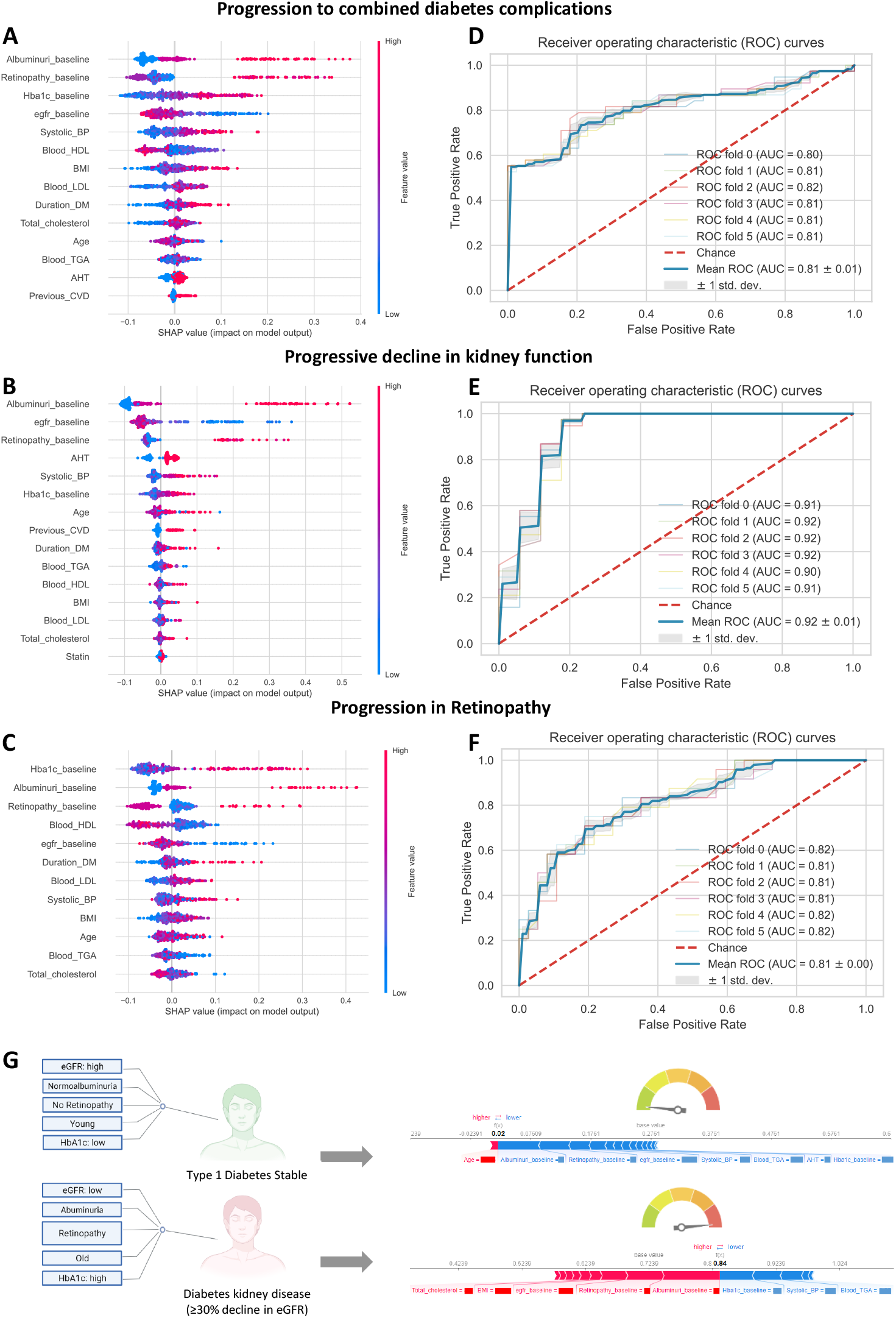
Models based clinical features. (**A-C**) Subset of a dot plot showing the directional mean absolute SHAP values of various features (x axis) computed from five-fold cross-validation models that predict metabolite levels (y axis) using clinical data. Positive and negative SHAP values represent positive and negative impact on the predicted risk of progression to combined DCs (**A**), ≥30% decline in eGFR (**B**), and retinopathy (**C**), respectively in the validation sets. Positive (negative) SHAP values indicate that higher (lower) feature values lead, on average, to higher predicted values. Each plot is made up of individual points from the validation dataset with a higher value being red and a lower value being blue. (**D-F**) AUROC, mean and SD of the result from model based on the main predictors. The 50 AUROC values for combined diabetes complications, diabetic kidney disease, and diabetic retinopathy obtained in MC had a mean and SD of 0.75±0.16, 0.75±0.14, and 0.96±0.25, respectively (not shown). (**G**) Force plots showing effect of SHAP values at the individual level performance of randomly predicted outputs (type 1 diabetes stable and type 1 diabetes with progression to ≥30% eGFR decline). Features in red show risk factors pushing up the overall probability while blue are protective factors. Feature labels are: **Retinopathy_baseline** (1= None apparent, 2= Mild non-proliferative, 3= Moderate non-proliferative); **Albuminuri_baseline** (1=normoalbuminuria, 2=microalbuminuria, 3=macroalbuminuria); **Previus_CVD** (1=yes); **AHT** (1=yes); **Statin** (1=yes).

**Fig. 3.**
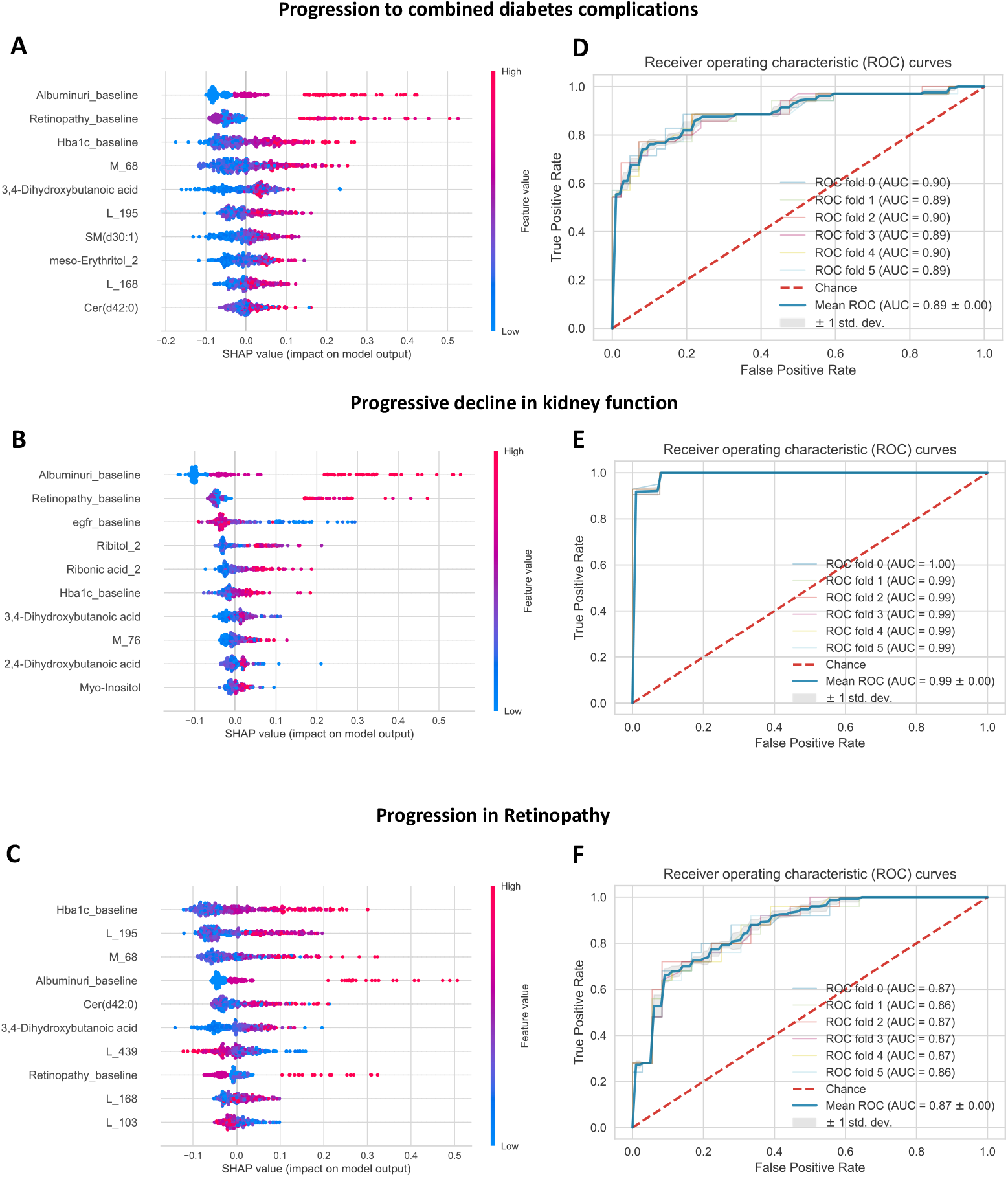
Models based clinical and omics features. (**A-C**) Subset of a dot plot showing the directional mean absolute SHAP values of various features (x axis) computed from five-fold cross-validation models that predict metabolite levels (y axis) using clinical data. Positive and negative SHAP values represent positive and negative impact on the predicted risk of progression to combined diabetes complications (**A**), ≥30% decline in eGFR (**B**), and retinopathy (**C**), respectively in the validation sets. Positive (negative) SHAP values indicate that higher (lower) feature values lead, on average, to higher predicted values. Each plot is made up of individual points from the validation dataset with a higher value being red and a lower value being blue. Shown are the top features by maximum mean absolute SHAP values across all clinical data. (**D-F**) AUROC, mean and SD result from model based on the main predictors. The 50 AUROC values for combined diabetes complications, diabetic kidney disease, and diabetic retinopathy obtained in MC had a mean and SD of 0.81± 0.10, 0.96±0.06, and 0.79±0.16, respectively (not shown). Feature labels are: **Retinopathy_baseline** (1= Noneapparent, 2= Mild non-proliferative, 3= Moderate non-proliferative); **Albuminuri_baseline** (1=normoalbuminuria, 2=microalbuminuria, 3=macroalbuminuria).

#### 3.3.1 Clinical-based biomarkers in the prediction of diabetes complications

Overall, 190 participants (49 %) experienced any progression of retinopathy and/or ≥ 30% decline in eGFR from baseline to follow-up. The final model for combined DCs selected through the SHAP method included 14 out of the initial 17 clinical baseline variables: albuminuria, mild degree of retinopathy, HbA_1c_, eGFR, systolic BP, HDL-cholesterol, BMI, LDL-cholesterol, diabetes duration, total cholesterol, age, total triglycerides, antihypertensive therapy (AHT) and previous cardiovascular disease (CVD) (**Figure 2.A**). Smoking, gender and statin remained unincluded. Using five-fold cross-validation for discrimination on SHAP selected features, the mean AUROC was 0.81 (95% CI 0.687;0.893) in the validation set with an accuracy of 0.81, precision of 0.79, F1-score of 0.80, and recall of 0.81 (**Figure 2.D**). The 50 AUROC values obtained in MC had a mean of 0.75, and a standard deviation of 0.16.

A total of 79 participants (21 %) experienced a progressive decline of ≥30 % in eGFR. The optimal DKD model included 15 clinical baseline variables: albuminuria, eGFR, mild degree of retinopathy, AHT, systolic blood pressure, HbA_1c_, age, previous CVD, diabetes duration, total triglycerides, HDL-cholesterol, BMI, LDL-cholesterol, total cholesterol, and statin, excluding smoking and gender (**Figure 2.B**). Smoking and gender were not included in the optimal model. AUROC for the DKD model was 0.92 (95% CI 0.857;0.995) with an accuracy of 0.95, precision of 1.00, F1-score of 0.89, and recall of 0.80 (**Figure 2.E**). The 50 AUROC values obtained in MC had a mean of 0.96, and a standard deviation of 0.25. An individual decision tree for classifying the ≥ 30% eGFR decline model based clinical dataset is shown in Fig. 4.A.

**Fig. 4.**
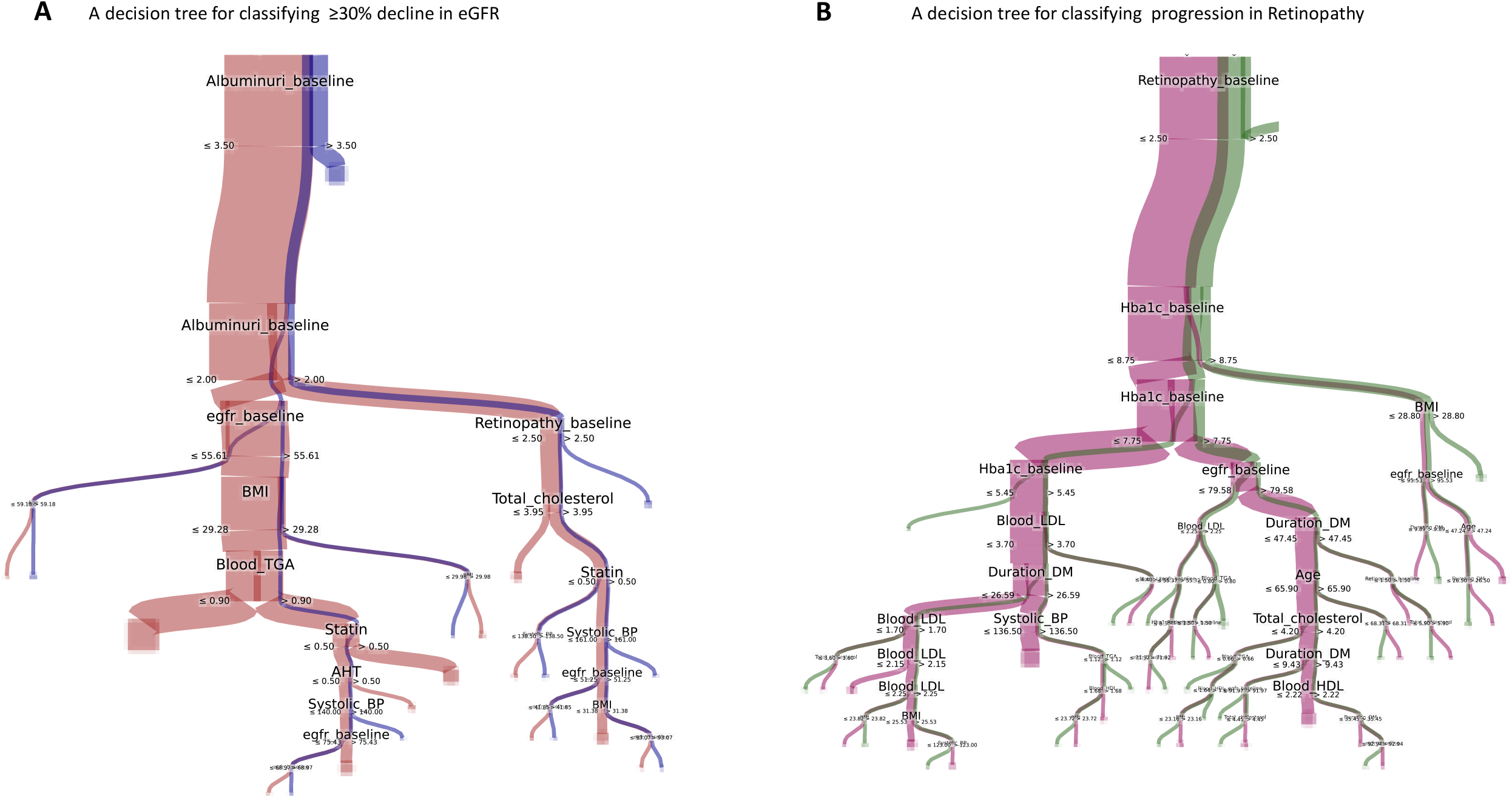
Visualization of random forest models based clinical features. **A**, Graph showing an example of a tree computed using random forest. It is one example out of the 500 computed decision trees for classifying the clinic dataset for predicting ≥30 % decline in eGFR. **B**, an example of one out of the 500 decision trees for classifying the clinic dataset for predicting retinopathy. Each class is represented by a color (brown and blue in A, and purple and green in B), the width of the link represents the number items flowing from one node to the other.

A total of 111 participants (28.83%) experienced any progression of retinopathy. The best model derived from RF algorithm for retinopathy included 12 clinical baseline variables: HbA_1c_, albuminuria, mild degree of retinopathy, HDL-cholesterol, eGFR, diabetes duration, LDL-cholesterol, systolic BP, BMI, age, total cholesterol, total triglycerides, and total cholesterol (**Figure 2.C**). Smoking, gender, statin, AHT, and previous CVD were not included in the optimal model. The mean AUROC for the retinopathy model was 0.81 (95% CI 0.754;0.958) with an accuracy of 0.75, precision of 0.73, F1-score of 0.59, and recall of 0.50 (**Figure 2.F**). The 50 AUROC values obtained in MC had a mean of 0.75, and a standard deviation of 0.14. An individual decision tree for classifying the retinopathy model based clinical dataset is shown in Fig. 4. B.

Feature importance and personalized individual risk predictions of DKD, progressors versus non-progressors, were examined further (**Figure 2.G)**. The first force plot shows a stable individual without progression to DCs and correctly predicted as a non-progressor by the model: the predicted probability of progression was 2 %. The second force plot shows an individual correctly predicted as progressor with the probability of 84 %. In more detail, the SHAP values of individual participants emphasize variables that most strongly contribute to the prediction, with red and blue colors, respectively, indicating risk factors and protective factors. For instance, with the second individual predicted as DKD progressor, albuminuria, mild degree of retinopathy, and eGFR played an important role in the prediction: albuminuria was the most important risk factor as determined by the color (red) and the length of the respective bar. In contrast, the first individual was predicted to remain free of DKD based on young age, normo-albuminuria, no retinopathy, and a relatively high eGFR, all contributing to the very low probability, 2 %, of DKD progression.

#### 3.3.2 Omics and clinical profile-based biomarkers in the prediction of diabetes complications

The optimal model for any progression in DCs was obtained by combining three clinical baseline variables -- albuminuria, mild degree of retinopathy and HbA_1c_ -- with seven metabolites -- 3,4-Dihydroxybutanoic acid, SM(d30:1), meso-Erythritol, Cer(d42:0), one unidentified metabolite and two unidentified lipid species (**Figure 3.A**). This final model with SHAP-selected clinical variables and omics features had a mean AUROC of 0.89 (95% CI 0.818;0.966), accuracy of 0.83, precision of 0.90, F1-score of 0.81, and recall of 0.73 in the validation set (**Figure 3.D**). The 50 AUROC values obtained in MC had a mean of 0.81, and a standard deviation of 0.10.

The best model for DKD was obtained by combining four clinical baseline variables: albuminuria, mild degree of retinopathy, eGFR and HbA1c, and two ketones and three sugar derivatives: 3,4-dihydroxybutanoic acid, 2,4-dihydroxybutanoic acid, ribitol, ribonic acid, myo-inositol, and one unidentified metabolite (**Figure 3.B**). The model demonstrated an excellent performance, with mean AUROC=0.99 (95% CI 0.876;0.997), accuracy of 0.98, precision of 1.00, F1-score of 0.96, and recall of 0.92 (**Figure 3.E**). The 50 AUROC values obtained in MC had a mean of 0.96, and a standard deviation of 0.06. The best performing model for DR was based on seven metabolites: Cer(d42:0), 3,4-Dihydroxybutanoic acid, the same unidentified metabolite as in the model above, and four unidentified lipid species together with HbA1c, albuminuria, and mild degree of retinopathy (**Figure 3.C**). The mean AUROC was 0.87 (95% CI 0.781;0.996) with an accuracy of 0.80, precision of 0.68, F1-score of 0.71, and recall of 0.75 (**Figure 2.F**). The 50 AUROC values obtained in MC had a mean of 0.79, and a standard deviation of 0.16.

Both models with and without adding blood small-molecules provided a good discrimination to predict DCs. The recall of the models with clinical variables indicates that 80 % and 50 %, respectively, were correctly identified as progressors to ≥30 % decline in eGFR and any retinopathy. In models with, both, clinical variables and small molecules, 92 % and 80 % were correctly identified as progressors to ≥30 % decline in eGFR and any retinopathy, respectively.

## 4. Discussion

In the present study, we developed high-performing prediction models with random forest ML algorithms utilizing clinical risk factors and omics profiles from plasma samples of persons with type 1 diabetes. Our objective was to predict progression of diabetic kidney disease, defined as ≥30 % decline in eGFR, and retinopathy defined as progression in retinopathy severity over 5 years.

Using only clinical risk factors for training the models, AUROC of 0.81, 0.92, and 0.81 were obtained for combined DCs, DKD, and DR respectively (**Figure 2.D-F**). The models based on the clinical risk profile accurately predicted the future progression of DCs in individuals with type 1 diabetes. Moreover, prediction improved by the inclusion of blood biomarkers from the omics data (**Figure 3.D-F**). Including a molecular profile to the predictive panel may be useful for the implementation of detailed personalized medicine tools in the clinic. However, molecular panels need further investigation, including the testing of clinical utility with clinical trials (13).

The models with clinical risk factors were obtained with routinely collected data (such as HbA_1c_, albuminuria and eGFR) all known risk factors of microvascular complications in diabetes (4, 30). Albuminuria, eGFR and retinopathy status at baseline were the main predictors for ≥30 % decline in eGFR progression. Similarly, HbA_1c_, albuminuria, and retinopathy status were main predictors for progression to DR.

In our models, baseline DR was one of the top three variables of importance for predicting future DKD. The association of diabetes nephropathy and DR has been addressed in several previous studies (31-34), confirming the plausibility of the three main predictors over other clinical factors.

Overall, we identified eight small biomolecules from the models with clinical risk factors and blood-derived molecular data that were strongly predictive of DCs. The metabolite signature to predict ≥30 % decline in eGFR included two short chain ketones (3,4-dihydroxybutanoic acid and 2,4-dihydroxybutanoic acid) and three sugar derivatives (myo-inositol, ribitol, and ribonic acid). Ribitol and ribonic acid were the main metabolite predictors (**Figure 3.C**). Ribonic acid and ribitol are sugar acid derivatives from ribose and are involved in the pentose phosphate pathway. In accordance with the present results, elevated levels of ribitol are associated with retinal cell apoptosis in DR (35). Moreover, elevated levels of ribitol and myo-inositol in chronic kidney disease stages 3-5 have been reported (36).

Myo-inositol is involved in inositol metabolism and is primarily synthesized in the kidneys at a rate of a few grams per day in humans. The overexpression of myo-inositol oxygenase has been suggested to drive the progression of renal tubulointerstitial injury in a mouse model of diabetes (37). Previous results in type 2 diabetes also showed that higher levels of myo-inositol were associated with a higher risk of end stage renal disease (38). In the present study, we show that higher levels of myo-inositol were predictive of ≥30 % decline in eGFR (**Figure 3.B**).

The metabolite signature to predict retinopathy progression included 3,4-dihydroxybutanoic acid and a saturated ceramide (Cer(d42:0)). An earlier metabolomics study by Chen et al. identified 3,4-dihydroxybutanoic acid as a novel biomarker for DR (39). Ceramides are sphingolipids, which are active in cell-signaling processes, also associated with the pathogenesis of diabetes, insulin resistance and heart disease (40, 41). In the present study, DR progressors showed increased levels of Cer(d42:0) and 3,4-dihydroxybutanoic acid at baseline when compared with non-progressors with diabetes (**Figure 3.B**).

Evidence from the present study shows that prediction models based on variables routinely collected in the clinic can be excellent predictors of individual prognosis. The results suggest that the measurement of relevant biomolecules from the circulation can further improve the accuracy of these predictions.

Furthermore, we argue that biomolecules may be necessary for a more fine-grained understanding and prediction of complications, which will be necessary for personalized medicine in practice.

In previous studies from other cohorts we have seen other omics-based markers associated to progression of kidney disease using urinary proteomics and, in the future,, it will be interesting to see if the combination of omics panels from two biofluids can improve prediction (42).

### Strengths and limitations

Our study benefits from a large and comprehensive dataset with a good representation of individuals who progressed to two different DCs. This allowed us to test, both, routinely collected clinical data as well as molecules that are measured with advanced mass-spectrometry.

The ML models with and without omics were robust with stable performance across the cross-validation. Yet, a limitation is that the study was based on a single cohort, although this was attenuated in part by the model being validated on unseen data representing twenty percent of the cohort. Therefore, replication in a clinical trial will be of substantial interest and necessary for implementing this tool for clinical decision making (9). Except for the outcomes of DCs, the predictor data were restricted to a snapshot baseline profile. Therefore, longitudinal tracking of molecular data could contribute to more accurate and robust prediction.

According to a newly-published report from the American Diabetes Association (ADA) and European Association for the Study of Diabetes (EASD) (1), advanced data and algorithms are expected to contribute to better clinical decision making. Predicting DCs before their onset is very challenging in real-world clinical practice, and early detection can have major implications on the quality and length of life. Our aim is to further increase the understanding of how individuals with diabetes progress towards harmful complications. We believe that ML-based high-performing predictive models will support clinicians in these challenging decisions.

In conclusion, we have demonstrated that ML algorithms using traditional risk factors can successfully predict future progression of DCs in type 1 diabetes. The inclusion of omics data further improved the predictions. We believe that with further development and validation, the prediction models presented here have the potential for early detection of complications, thus enabling appropriate interventions to be taken to prevent further progression of these complications.

## Data Availability

Data are available on request for researchers who have acquired the required legal permissions from the Danish data protection agency. Requests to access the datasets should be directed to PR.

## Author Contributions

N.A., C.L-Q., P.R., and D.M. conceived the study concept and design. T.S. and P.H. contributed to data curation. N.A., S.K., D.M., and C.L-Q. contributed to development of methodology. N.A. and S.K. performed the data analysis. N.A. drafted the manuscript. All authors critically revised the manuscript and approved the final version. P.R., F.P., and C.L-Q. contributed to funding acquisition. C.L-Q. is guarantor of this work and, as such, had full access to all the data in the study and takes responsibility for the integrity of the data and the accuracy of the analysis.

## Acknowledgments

We thank all participants of the study. We thank the laboratory technicians at Steno Diabetes Center Copenhagen, Gentofte, Denmark, for their excellent technical assistance. We acknowledge the support from the Novo Nordisk Foundation Challenge grant PROTON (Personalized treatment of diabetic nephropathy) NNF14OC0013659.

## Code availability

The ML script has been deposited in a GitHub repository (available here: https://git.io/JD5A2) to improve reproducibility and transparency.

## Ethics declarations

The study involving human participants were approved by The Ethics Committee E, Region Hovedstaden, Denmark. The participants /patients provided their written informed consent to participate in this study.

